# Does Copper treating of commonly touched surfaces reduce healthcare acquired infections? A Systematic Review and meta-analysis

**DOI:** 10.1101/2020.05.21.20109447

**Authors:** Loai Albarqouni, Oyuka Byambasuren, Justin Clark, Anna Mae Scott, David Looke, Paul Glasziou

## Abstract

**Background:** Healthcare acquired infections (HAIs) cause substantial morbidity and mortality. Copper appears to have strong viricidal properties under laboratory conditions.

**Aim:** We conducted a systematic review to examine the potential effect of copper treating of commonly touched surfaces in healthcare facilities.

**Methods:** We included controlled trials comparing the effect of copper-treated surfaces (furniture or bed linens) in hospital rooms versus standard rooms on hospital acquired infections (HAIs). Two reviewers independently screened retrieved articles, extracted data, and assessed the risk of bias of included studies. The primary outcome was the occurrence of healthcare acquired infections.

**Findings:** We screened 638 records; 7 studies comprising 12362 patients were included. All of included studies were judged to be at high risk in ≥2 of the 7 domains of bias. All 7 included studies reported the effect of copper-treated surfaces HAIs. Overall, we found low quality evidence of a potential clinical importance that copper-treated hard surfaces and/or bed linens and clothes reduced healthcare acquired infections by 27% (RR 0.73; 95% CI 0.57 to 0.94).

**Conclusion:** Given the clinical and economic costs of healthcare acquired infections, the potentially protective effect of copper-treated surfaces appears important. The current evidence is insufficient to make a strong positive recommendation. However, it would appear worthwhile and urgent to conduct larger-scale publicly funded clinical trials of the impact of copper coating.

## INTRODUCTION

Finding effective and sustainable ways of reducing pathogen transmission is important for all epidemics but particularly urgent in the current SARS coronavirus 2 (SARS-CoV-2) pandemic^1^. Though the exact relative importance of different modes of transmission is currently unknown there appears to be three main avenues, namely direct aerosol, contact with fomites, and the most controversial, airborne transmission^2^. Reducing the incidence of infections will require addressing all modes of transmission. While social distancing is widely promoted it may not completely prevent fomite transmission if common objects such as doorhandles, stair banisters, table surfaces, utensils or taps are contaminated^3^.

While the survival of viruses such as SARS-CoV-2 on surfaces varies with temperature, humidity and many other factors. Several studies suggest viable virus can remain for several days on some materials with a 10-fold reduction per day in viral load^4^. Transmission from an infected person to a new person via a fomite such as a door handle, tap or even money (and subsequent self-inoculation via nose, eyes or mouth), is suspected to be significant, second only to direct droplet transmission^5^. For example an analysis of an outbreak in a Chinese shopping centre found that transmission occurred from the initial infected group on the seventh floor to staff on other floors with whom they had had no contact but had shared elevators and restrooms^6^. In this case, transmission may have been through fomite contamination of lift buttons, handles, or taps, or potentially airborne transmission via enclosed spaces, although this is controversial. Cleaning shared surfaces is one proposed preventive mechanism but would require frequent and extensive cleaning.

Copper appears to have strong viricidal properties and substantially reduces the duration of viral viability on surfaces from days to 30 to 60 minutes under laboratory conditions^7^. The inactivation property of copper has been demonstrated for both Norovirus and for Coronavirus species with inactivation occurring in less than 60 minutes^7^. Inactivation also occurs on copper alloys and the activity appears directly proportional to the percentage of copper present in the alloy. This property has led researchers to examine the potential for copperplating of common surfaces to reduce healthcare acquired infections with multi-resistant bacteria as well as viruses with attempts to copperplate common shared surfaces in hospital wards. These include surfaces such as bedrails, door handles, table surfaces, as well as soft textiles such as bed linen, patient gowns and towels. If coating of commonly touched surfaces in hospital rooms could reduce healthcare acquired infections, the impact could be substantial in both health and economic terms. Therefore, we aimed to examine the potential of copper coating of common shared surfaces in hospitals. We aimed to find all controlled trials which had compared copper-treated surfaces in hospital rooms or items with standard rooms or items.

## METHODS

We aimed to find, appraise, and synthesize eligible studies that have compared the effect of copper-treated hospital rooms versus standard rooms on healthcare acquired infections. This systematic review is reported following the Preferred Reporting Items for Systematic Reviews and MetaAnalyses (PRISMA) statement and the review protocol was prospectively developed^8^.

### Eligibility criteria

*Participants*. We included studies of patients of any age and with any condition in acute and longterm care settings (including intensive care units, rehabilitation centres, and aged-care facilities).

*Interventions*. We included studies that evaluated interventions involving copper (or copper alloy) surfaced rooms or objects in patient care rooms/spaces. We expanded the intervention to include studies evaluated copper-treated soft textiles such as bed linens, clothes, and gowns as sufficient data was available.

*Comparators*. We included studies with any comparator, as long as it did *not* involve the use of copper or copper alloy surfaces.

*Outcomes (primary, secondary)*. The primary outcome was the incidence of healthcare acquired infection (e.g. bacterial or viral infections – *not* colonisations) in patients. The secondary outcomes were the incidence of deaths and any skin reactions in patients, and any healthcare acquired infection (e.g. bacterial or viral) in hospital staff and visitors. We excluded studies that only reported the rate of colonisations (not infections).

*Study design*. We included randomised and pseudo-randomised (e.g. alternate allocation) controlled trials.

### Search strategies to identify studies

#### Database search strings

We searched PubMed, Cochrane CENTRAL and Embase from inception until 25 March 2020. We designed a search string in PubMed that included the following concepts: Copper AND infections AND healthcare facility AND controlled trial. The PubMed search string was translated using the Polyglot Search Translator^9^ and run in the other two databases **(Appendix 1)**

#### Restriction on publication type

No restrictions by language or publication date were imposed. We included publications that were published in full; publications available as abstract only (e.g. conference abstract) were included if they had a clinical trial registry record, or other public report, with the additional information required for inclusion. We excluded publications available as abstract only (e.g. conference abstract) with no additional information available.

#### Other searches

On 26 March 2020 we conducted a backwards (cited) and forwards (citing) citation analysis in Scopus on the included studies identified by the database searches. These were screened against the inclusion criteria. Clinical trial registries were searched on 25 March 2020 via Cochrane CENTRAL, which includes the WHO ICTRP and clinicaltrials.gov.

### Study selection and screening

Two authors (LA, OB) independently screened the titles and abstracts for inclusion against the inclusion criteria. One author (JC) retrieved full-texts, and two authors (LA, OB) screened the full-texts for inclusion. Any disagreements were resolved by discussion, or reference to a third author (PG). The selection process was recorded in sufficient detail to complete a PRISMA flow diagram (see **Figure 1**) and a list of excluded full-text articles with reasons for exclusions (see **Appendix 2**).

**Figure 1.**
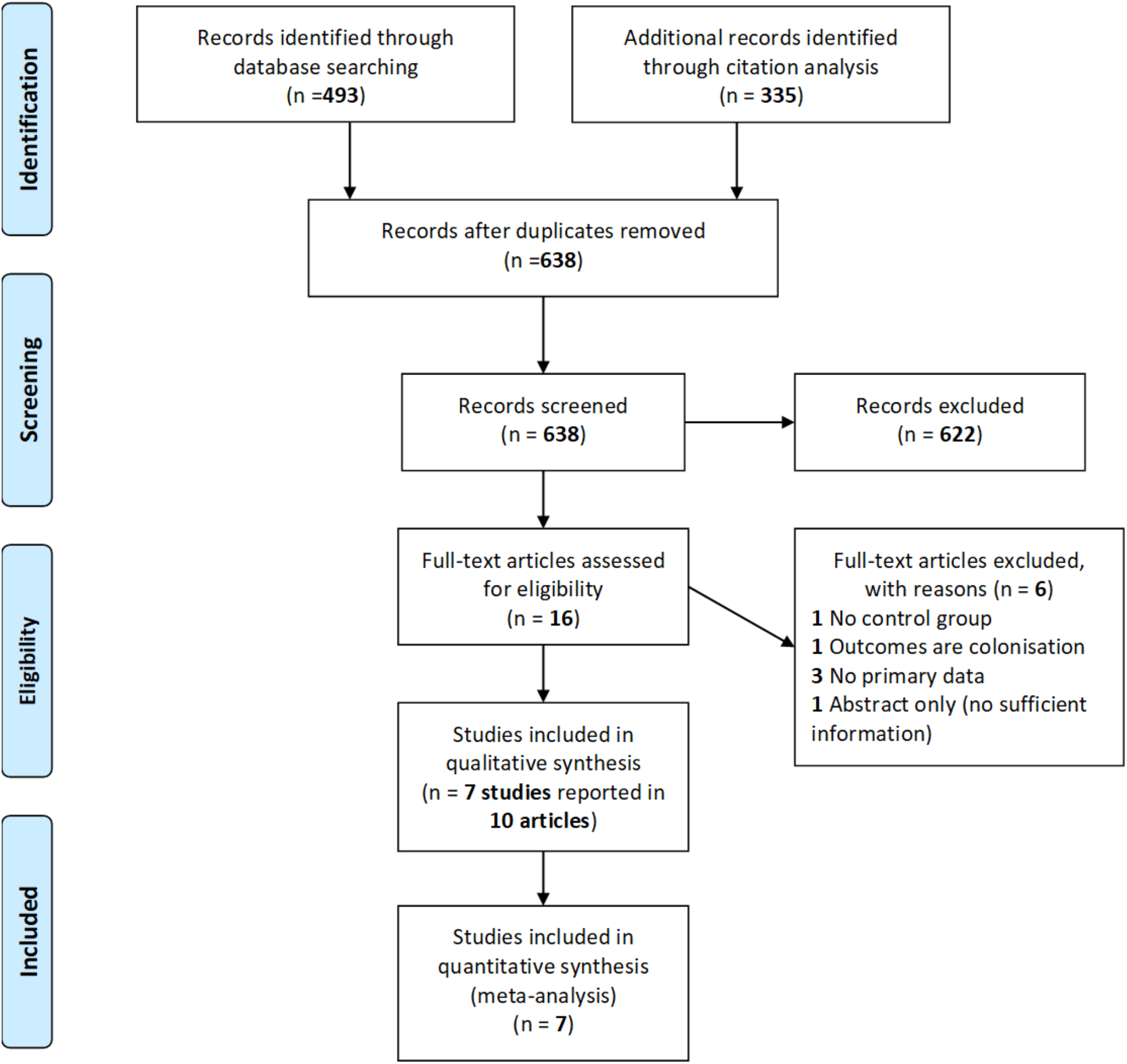
PRISMA flow diagram of included articles

### Data extraction

We used a data extraction form for study characteristics and outcome data, which was piloted on two studies in the review. Two authors (LA, OB) extracted the following data from included studies:

1. Methods: study authors, location, study design, duration of study, duration of follow-up
2. Participants: N, age (mean or median; range), gender, diagnosis or infection type at admission, ward or room type admitted to (e.g. intensive care, acute care, long-term care)
3. Interventions and comparators: type of copper coating (e.g. copper percentage in the alloy), type of surfaces covered by copper/copper alloy (e.g. bed controls, tables, etc.), type of comparator, average duration of stay in the room.
4. Outcomes: primary and secondary outcomes: incidence of healthcare acquired infections (e.g. bacterial or viral infections) in patients (*primary*), or hospital staff or visitors (*secondary*), and the number of deaths (secondary) and skin reactions (*secondary*).

### Assessment of risk of bias in included studies

Two review authors (LA, OB) independently assessed the risk of bias for each included study using the Risk of Bias Tool 1, as outlined on the *Cochrane Handbook^10^*. All disagreements were resolved by discussion or by referring to a third author (PG). The following domains were assessed:

1. Random sequence generation
2. Allocation concealment
3. Blinding of participants and personnel
4. Blinding of outcome assessment
5. Incomplete outcome data
6. Selective outcome reporting
7. Other bias (focusing on potential biases due to funding or conflict of interest).

Each potential source of bias was graded as low, high or unclear, and each judgement was supported by a quote from the relevant trial.

### Measurement of effect and data synthesis

We used risk ratios or rate ratios for dichotomous outcomes – risk ratios for results reporting the number of patients with an event, and rate ratios for the results reporting the number of events only. We undertook meta-analyses only when meaningful (when ≥2 studies or comparisons reported the same outcome); anticipating considerable heterogeneity, we used a random effects model. We used *Review Manger 5* to calculate the intervention effect.

### Assessment of heterogeneity and reporting biases

We used the I^2^ statistic to measure heterogeneity among the included trials. Because we included fewer than 10 trials, we did not create a funnel plot.

### Dealing with missing data

We contacted investigators or study sponsors to provide missing data.

### Subgroup and sensitivity analyses

We planned to do a subgroup analysis by type of infection/patient and a sensitivity analysis by including versus excluding studies at high risk of bias, however, due to a low number of included studies, these analyses were not done.

## RESULTS

We screened 638 titles and abstracts and assessed 16 full-text articles for inclusion. After excluding 6 articles, we included 10 articles pertaining to 7 studies^11–20^. We also identified 5 relevant clinical trial registries (2 for studies already identified and included and 3 registries for studies that have not been published). **Figure 1** shows PRISMA flow diagram of studies. Excluded full-text articles are presented in **Appendix 2** with reasons for exclusion.

### Characteristics of included studies

We included 7 controlled studies, which enrolled a total of 12,362 participants^12,13,15,16,18–20^. Included studies were conducted in the last decade in the USA (n=3 studies^13,16,18^), Chile (n=2 studies^15,19^), France (n=1 study^20^), and Israel (n=1 study^12^). Three of the studies were set in adult ICUs^13,16,18^, one in paediatric ICU^19^, one in aged care facility^20^, one in acute care ward^15^, and one in long-term care for ventilator dependent patients^12^. Duration of the studies ranged from 7 to 16 months.

Four of the included studies evaluated the effect of copper coating of commonly touched hard surfaces such as bed rails and tables, IV poles, door handles and taps on healthcare acquired infections^15,16,19,20^. Two studied copper-treated linens (bedding, patient gowns and towels)^12,13^ and one included both hard surfaces and linens^18^. All included studies reported the effect of copper on healthcare acquired infections in patients (i.e. primary outcome); none reported the effect on hospital staffs or visitors (i.e. secondary outcome).

### Risk of bias assessment

All of the 7 included studies were judged to be at high risk in two or more of the domains of bias. Of the 7 included studies, 5 were judged to be at high or unclear risk for selection bias (either random sequence generation or allocation concealment). All of included studies were judged to be at high or unclear risk in blinding of participants or personnel and conflict of interest (recorded as “other risk of bias”). All of included studies were judged to be at low risk in attrition bias (i.e. incomplete outcome data) and reporting bias (i.e. selective reporting).

### Effects of copper-treated surfaces

#### Healthcare acquired infections (HAIs)

All 7 included studies reported the effect of copper-treated surfaces on healthcare acquired infections. Overall, we found that copper-treated hard surfaces and/or bed linens and clothes reduced healthcare acquired infections by 27% (RR 0.73; 95% CI 0.57 to 0.94) (**Figure 2**).

**Figure 2:**
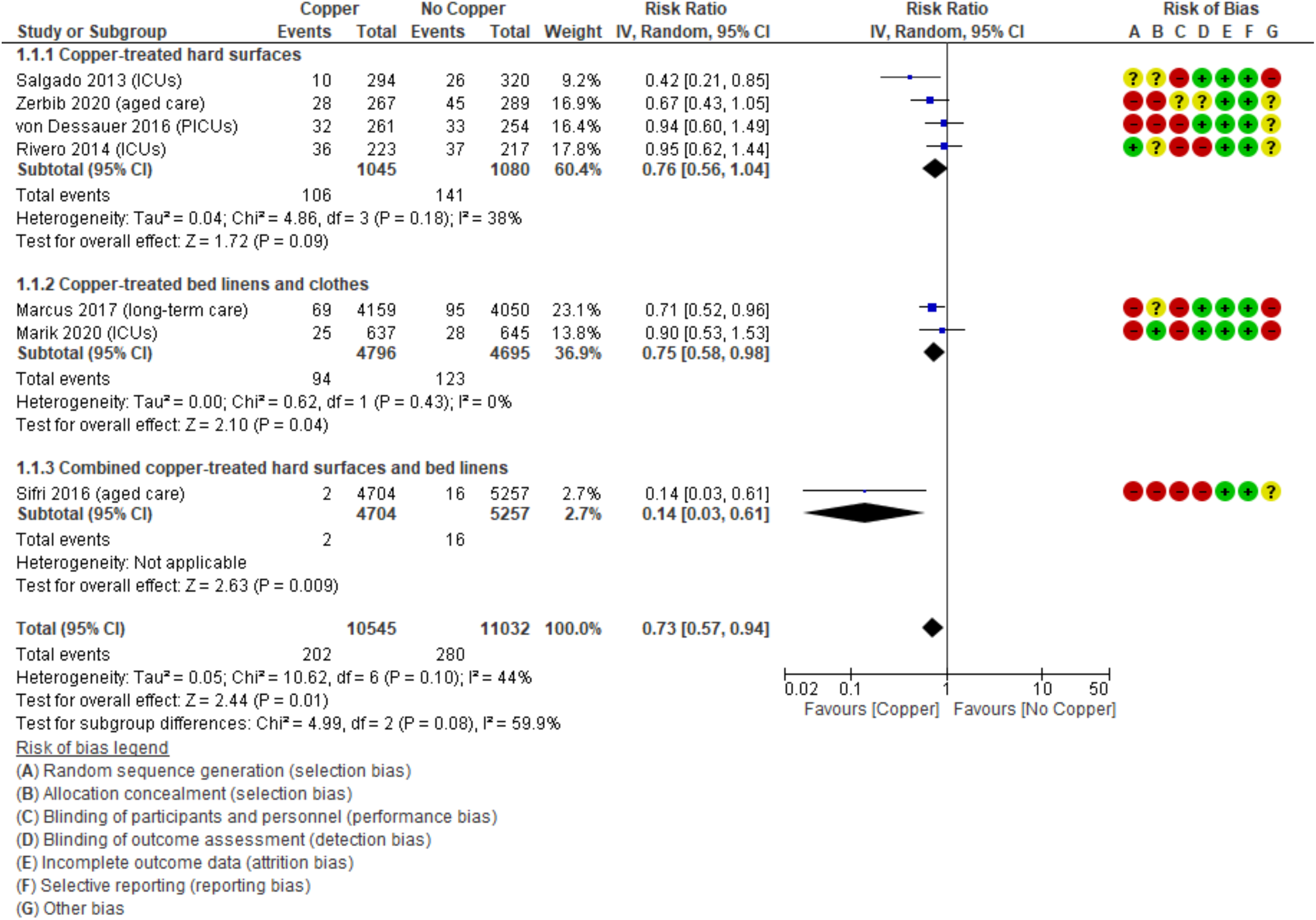
Forest plot of healthcare acquired infections in copper treated surfaces versus no copper. *Abbreviation: ICUs, Intensive Care Units; PICUs, Paediatric ICUs*.

##### Copper-treated hard surfaces (4 studies)

We identified 4 studies (2125 participants) that evaluated the effect of copper-treated hard surfaces on healthcare acquired infections^15,16,19,20^. There was no statistically significant reduction in HAIs among participants hospitalised in facilities with copper-treated surfaces compared to no copper (RR 0.76, 95% CI 0.56 to 1.04; I^2^=38%).

##### Copper-treated bed linens and clothes (2 studies)

We identified 2 studies (276 participants) that evaluated the effect of copper-treated bed linens and clothes on HAIs^12,13^. We observed a statistically significant 25% relative reduction in HAIs among participants hospitalised in facilities with copper-treated bed linens and clothes compared to no copper (RR 0.75, 95% CI 0.58 to 0.98; I^2^=0%).

##### Combined copper treated hard surfaces and bed linens and clothes (1 study)

A single study of 9,961 participants evaluated the combined effect of both copper-treated hard surfaces and bed linens and clothes on HAIs^18^. A statistically significant 86% relative reduction in HAIs was observed among participants hospitalised in facilities with copper-treated surfaces compared to no copper (RR 0.14, 95% CI 0.0.03 to 0.61).

### Mortality

Of the 7 included studies, 3 studies (included a total of 1,569 participants) reported the effect of copper-treated hard surfaces on mortality^15,16,19^. There was no statistically significant difference in mortality between participants hospitalised in facilities treated with copper compared to no copper (RR 1.06, 95% CI 0.83 to 1.36) (**Figure 3**).

**Figure 3:**
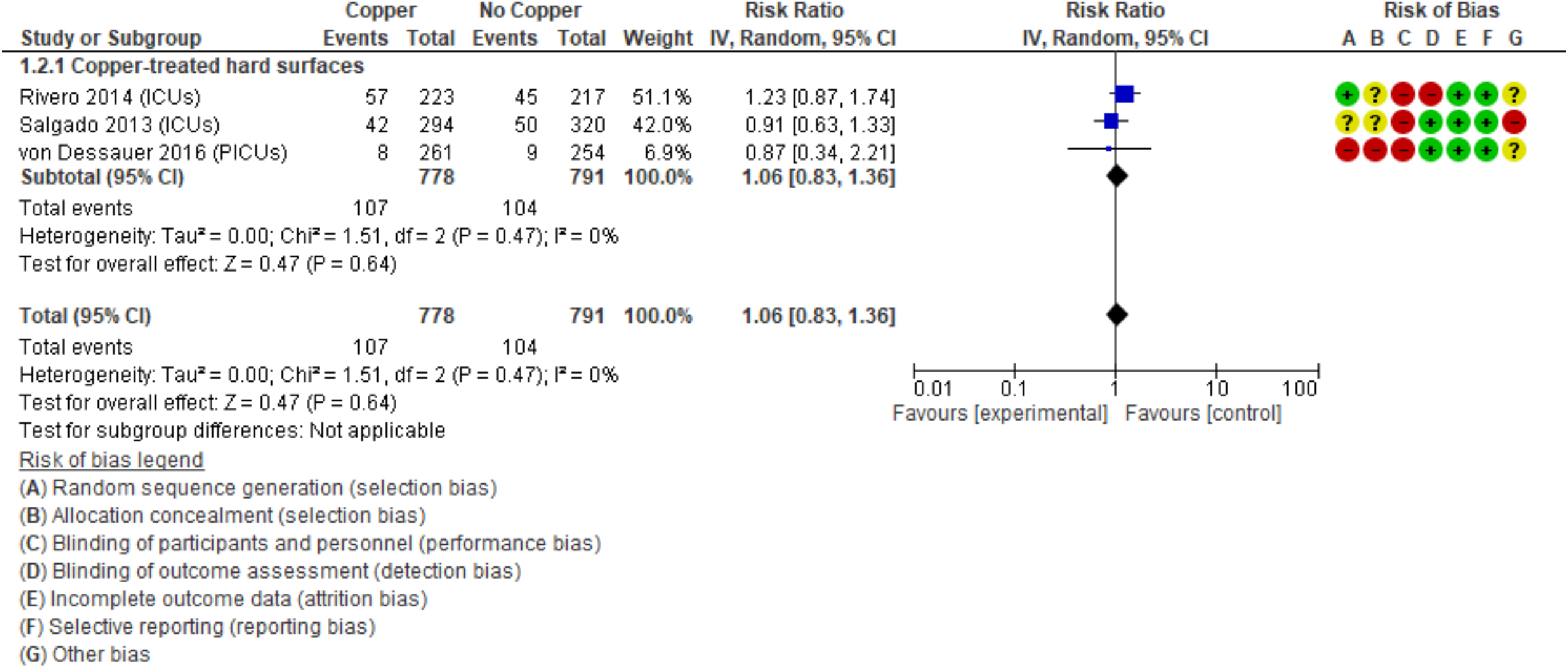
Forest plot of mortality in copper treated surfaces versus no copper. *Abbreviation: ICUs, Intensive Care Units; PICUs, Paediatric ICUs*.

**Table 1.** Characteristics of included studies (n=7)

| Study ID, year and country | Study type and duration | Participants, Setting | Intervention | Control | Primary outcomes |
| --- | --- | --- | --- | --- | --- |
| **Salgado 2013 (USA)** | Double-blind RCT, 11 months | 614 adult ICU patients (60.4 yrs) | *Copper-treated surfaces*<br>Copper covering of bed rails, overbed tables, intravenous poles, and arms of the visitor's chair the nurses' call button, computer mouse, bezel of touchscreen monitor, palm rest of laptop were different depending on the hospitals) | Regular ICU | Incident Rate of HAI and/or MRSA or VRE colonization |
| **Rivero 2014 (Chile)** | Controlled trial, 13 months | 440 adult ICU patients (51 yrs) | *Copper-treated surfaces*<br>C11000 copper alloy (99% copper) equivalent to approximately 80% of the areas most touched by patients (four bed rails, patient table and two IV poles) | Regular ICU | HAI, associated mortality, cost of antimicrobials |
| **von Dessauer 2016 (Chile)** | Non-randomized, unmasked, controlled clinical trial, 12 months | 65 paediatric ICU (1 yr) | *Copper-treated surfaces*<br>Copper covering of bed rails, bed rail levers, intravenous poles, sink handles, and the nurses' workstation | Regular PICU | Diagnosis of an HAI event associated with patient stay within the PICU or PIMCU |
| **Zerbib 2020 (France)** | Controlled trial, 16 months | 556 nursing home residents (85.4 yrs) | *Copper-treated surfaces*<br>438 door handles, 322 m of handrails, and 10 grab-bars in copper alloy (containing 90% copper) | Regular nursing home setting | Rates of infection during outbreak (5 cases in 4 days) |
| **Marcus 2017 (Israel)** | Double-blind, controlled cross-over, 7 months (2 x 3 months, separated by a 1-month washout period) | 112 ventilator-dependent patients in a long-term care hospital (69.8 vs 71.3 yrs) | *Copper-treated textiles*<br>Copper oxide-impregnated linen and hospital patients' clothes and towels. | Regular ICU | Antibiotic treatment initiation events (ATIEs), fever days, days of antibiotic treatment, and antibiotic defined daily dose (DDD) per 1,000 hospitalization days (HDs) |
| **Marik 2020 (USA)** | Prospective, cluster, cross-over, randomized control trial, 11 months (2 x 5 months separated by 2 weeks of wash-out) | 1282 adult ICU patients (60 yrs) | *Copper-treated textiles*<br>Copper-oxide-treated **linens** (top sheets, fitted sheets, pillowcases, under pads, wash cloths, towels, and patient gowns) | Regular ICU | HCAI |
| **Sifri 2016**<br>**(USA)** | Quasi-experimental study with a control group, 10 months | 9961 adult acute care patients (58.5 vs 60.5 yrs) | *Combined Copper treated textiles and surfaces*<br>16% copper oxide-impregnated composite countertops (sinks, vanities, desks, computer stations, soiled utility rooms, nurse workstations) and moulded surfaces (overbed tray tables, bedrails) and copper-impregnated woven linens (patient gowns, bedding, washcloths, towels, bath blankets, thermal blankets) | Regular ICU | Incidence Rate of hospital-onset infections (C difficile or MDRO) |

ICU – Intensive Care Unit/s; HAI -healthcare acquired infection/s; MRSA – methicillin-resistant Staphylococcus aureus; VRE - vancomycin-resistant Enterococcus; PICU – Paediatric Intensive Care Unit/s; PIMCU - intermediate pediatric care unit; HCAI - healthcare-associated infections; MDRO - multidrug resistant organisms;

### Skin reactions

Of the 7 included studies, 2 studies reported data on skin reactions^18,19^. von Dessauer et al did not observe any adverse events (i.e. skin or other allergic reactions) among any participants in either group^19^. Sifri et al reported that 10 (of 4707) patients hospitalised in copper-treated rooms developed skin rashes (9 were evaluated by a dermatologist and attributed to alternative aetiology and 1 was discharged before evaluation)^18^.

## DISCUSSION

We found seven controlled trials, which when combined suggest that copper surfacing or use in bed linen may have some effect on reducing healthcare acquired infections. The combined studies suggest a modest but potentially important effect.

There are several limitations to our findings. First, many of the studies were poorly reported, preventing a clear appraisal of the methods. Second, even when reporting was clear, the research methods often involved flaws in study design which might introduce bias. Finally, the small total numbers of infections meant that the confidence intervals around effects were wide, indicating considerable uncertainty in the size of any effect. The poor quality of reporting and methods, and small sizes of the studies would both downgrade the overall quality of the evidence, rating it - in GRADE terms - as low-quality evidence but of a potentially clinically important effect. In addition to these problems, many of the investigator teams had a conflict of interest with companies involved in copper use.

We report one difference between the protocol and the review: we had initially intended to include only studies of copper-plating of hard surfaces such as furniture. However, as several studies assessed the impact of copper-treating of textiles (clothing and/or bed linens) we broadened our inclusion criteria. This resulted in an inclusion of two clothing/linen-only studies^12,13^ and one study that assessed the impact of both furniture coating and Copper-impregnated textiles^18^.

The only previous systematic review we could identify was a 2017 report prepared by Cochrane Australia for Australia's National Health and Medical Research Council (NHMRC) which found 2 of these studies^16,19^, and concluded that “With only two non-randomised trials, both with uncertain results, it is not possible to draw conclusions from this evidence.” The three trials since then, plus two not identified in the 2017 review, have strengthened the body of evidence, but not sufficiently to be able to make strong recommendations.

Given the clinical and economic costs of healthcare acquired infections, the potential effect of copper coating appears important. We feel the current evidence is insufficient to make a positive recommendation. However, it would appear worthwhile and urgent to conduct larger-scale publicly funded clinical trials with clearly defined outcomes into the impact of copper coating. If such studies were to be funded, it would also be important to collect additional data such as the separation of bacterial and viral infections and measuring outcomes for healthcare workers as well particularly for viral infections.

## Data Availability

Extracted data are available on request to the corresponding author.

## Acknowledgments

None

## Contributions

LA, OB, JC, AMS, and PG designed the study. LA and OB screened articles, assessed study eligibility and quality, and extracted data. JC with the help of all study authors designed the search strategy. LA, PG, AMS wrote the first draft of the manuscript. All authors contributed to the interpretation and subsequent edits of the manuscript. LA is the guarantor.

## Data sharing

Extracted data are available on request to the corresponding author.

## Prior presentation

None

## Supporting Information

## APPENDIX 1 DATABASE SEARCH STRINGS

### PubMed

(“Copper”[Mesh] OR Copper[tiab])

AND

(“Infections”[Mesh] OR “Equipment Contamination”[Mesh] OR “Infection Control”[Mesh] OR “Cross Infection”[Mesh] OR Infection[tiab] OR Infections[tiab] OR Colonization[tiab])

AND

(“Health Facilities”[Mesh] OR Hospital[tiab] OR Hospitals[tiab] OR “Healthcare facility”[tiab] OR “Healthcare facilities”[tiab] OR “Intensive care”[tiab] OR “Intensive-care”[tiab] OR ICU[tiab] OR PICU[tiab] OR Ward[tiab] OR Wards[tiab])

AND

(Randomized controlled trial[pt] OR controlled clinical trial[pt] OR randomized[tiab] OR randomised[tiab] OR placebo[tiab] OR “drug therapy”[sh] OR randomly[tiab] OR trial[tiab] OR groups[tiab] OR Control[tiab] OR Controlled[tiab] OR Comparing[tiab] OR Compared[tiab])

NOT

(Animals[Mesh] not (Animals[Mesh] and Humans[Mesh]))

NOT

(“Case Reports”[pt] OR Editorial[pt] OR Letter[pt] OR “Comment”[pt] OR Meta-Analysis[pt] OR “Observational Study”[pt] OR “Systematic Review”[pt] OR “Case Report”[ti] OR “Case series”[ti] OR Meta-Analysis[ti] OR “Meta Analysis”[ti] OR “Systematic Review”[ti])

### Cochrane CENTRAL

([mh Copper] OR Copper:ti,ab)

AND

([mh Infections] OR [mh “Equipment Contamination”] OR [mh “Infection Control”] OR [mh “Cross Infection”] OR Infection:ti,ab OR Infections:ti,ab OR Colonization:ti,ab)

AND

([mh “Health Facilities”] OR Hospital:ti,ab OR Hospitals:ti,ab OR “Healthcare facility”:ti,ab OR “Healthcare facilities”:ti,ab OR “Intensive care”:ti,ab OR ICU:ti,ab OR PICU:ti,ab OR Ward:ti,ab OR Wards:ti,ab)

### Embase (via Elsevier)

